# Loneliness is associated with mentalizing and emotion recognition abilities in schizophrenia, but only in a cluster of patients with social cognitive deficits

**DOI:** 10.1101/2022.01.19.22269559

**Authors:** Ł. Okruszek, M. Jarkiewicz, A. Piejka, M. Chrustowicz, M. Krawczyk, A. Schudy, P.D. Harvey, D.L. Penn, K. Ludwig, M.F. Green, A. Pinkham

## Abstract

**Background:** Loneliness is a concern for patients with schizophrenia (SCZ). However, the correlates of loneliness in SCZ are unclear; thus, the aim of the study is to investigate neuro- and social cognitive (SC) mechanisms associated with loneliness in SCZ.

**Methods:** Data for the study were pooled from two cross-national samples (Poland/USA) and included 147 SCZ and 103 healthy controls (HC) overall. Data from clinical, neurocognitive, and SC assessments were examined as potential predictors of loneliness in HC and SCZ samples pooled across two sites. Furthermore, Latent Class Analysis (LCA) was used to cluster patients based on SC capacity. Next, the relationship between SC and loneliness was explored in each cluster of SCZ.

**Results:** SCZ reported higher levels of loneliness than HC. Loneliness was linked to increased negative and affective symptoms in patients. A negative association between loneliness and mentalizing and emotion recognition abilities was found in the patients with social-cognitive impairments, but not in those who performed at normative levels.

**Conclusions:** We have elucidated a novel mechanism which may explain previous inconsistent findings regarding the correlates of loneliness in SCZ. As decreased SC capacity may be linked with loneliness only in patients with observable SC impairments, SC heterogeneity in SCZ needs to be recognized while planning psychosocial interventions targeting loneliness in this group.

## Introduction

Social functioning deficits constitute one of the greatest therapeutic challenges in treatment of patients with schizophrenia (SCZ). Lower social functioning in SCZ compared to HC with large effect sizes is found consistently across all methods of clinical assessment, including self-report, clinical interview and performance-based measures (Schneider et al., 2017). The low frequency of social contacts observed in this group is a long-term predictor of more severe disability in SCZ (Siegrist, Millier, Amri, Aballéa, & Toumi, 2015). Moreover, clinical and cognitive predictors of social dysfunction experienced by individuals with schizophrenia have been extensively studied for the last two decades, with multiple studies supporting the notion that social cognition (SC) is a better predictor of community functioning than nonsocial cognition (Halverson et al., 2019).

Despite extensive research examining the mechanisms underlying reduced social functioning observed in individuals with schizophrenia, the issue of loneliness in this clinical group is relatively understudied. Loneliness is defined as a subjective mismatch between one’s existing and desired social relationships (Russell, Peplau, & Cutrona, 1980). Furthermore, loneliness is driven by one’s appraisal of his/her social relationships, rather than by objective characteristics of one’s social network, thus it is not synonymous with objective social disconnection, even though there is a correlation between loneliness and social disconnection (Cacioppo & Cacioppo, 2018). Importantly, findings from a large scale meta-analysis demonstrate that loneliness (26%) and objective social disconnection (29%) similarly increase mortality (Holt-Lunstad, Smith, Baker, Harris, & Stephenson, 2015).

As evidenced by two recent meta-analyses (Chau, Zhu, & So, 2019; Michalska da Rocha, Rhodes, Vasilopoulou, & Hutton, 2018), there is a significant relationship between loneliness and psychotic symptoms in clinical and nonclinical samples (Chau et al., 2019; Michalska da Rocha et al., 2018). Findings from a large-scale Australian cohort study showed that over 80% of SCZ experience loneliness and over one out of three patients (37.2%) identify loneliness and social disconnection as a top-most challenge in their everyday functioning (Stain et al., 2012). Loneliness in patients has been also linked to certain aspects of cardiometabolic health (Badcock et al., 2019). This finding is particularly important given the fact that cardiovascular disorders are the major contributor to increased mortality in individuals with schizophrenia (Ringen, Engh, Birkenaes, Dieset, & Andreassen, 2014).

Even though cognitive variables have not been included in the theoretical model of loneliness in individuals with psychosis (Lim, Gleeson, Alvarez-Jimenez, & Penn, 2018), increasing effort has been devoted to identifying such correlates of loneliness in SCZ. An association between loneliness and psychomotor speed was observed in a large cohort of Australian outpatients (Badcock et al., 2015), however no relationship between loneliness and executive functioning was found in a sample of 116 non-institutionalized patients (Eglit, Palmer, Martin, Tu, & Jeste, 2018).

Social cognitive correlates of loneliness have also been investigated due to the postulates of the evolutionary theory of loneliness formulated by Cacioppo (Cacioppo & Cacioppo, 2018). On one hand, increased vigilance to social threats could result in abnormal emotion recognition (Pinkham, Brensinger, Kohler, Gur, & Gur, 2011); while on the other hand, focusing solely on one’s view and reduced perspective taking could hinder one’s mentalizing capacity. An exploratory study by Trémeau, Antonius, Malaspina, Goff, & Javitt (2016) found a significant negative association between loneliness and both performance-based and self-report measures of SC in HC. However, a recent review of social information processing mechanisms in lonely individuals has found conflicting findings about the association between loneliness and emotion recognition in non-clinical populations (Spithoven, Bijttebier, & Goossens, 2017). Moreover, when we administered a battery of SC tests to non-clinical controls, no associations were found between lower-level social cue processing or higher-order mentalizing abilities and loneliness (Okruszek et al., 2021).

Interestingly, the connection between social cognition and loneliness may be more nuanced within clinical populations. For example, in the Trémeau et al. (2016) study, loneliness reported by SCZ participants was correlated with self-report, but not performance-based, measures of SC. However, there was no association between SC variables and loneliness reported by persons with schizophrenia in the study that was part of the multi-site US project [Social Cognition Psychometric Evaluation (SCOPE) project (Ludwig et al., 2020)]. However, based on the results of the study, Ludwig et al. (2020) proposed a testable hypothesis that the association between SC and loneliness may be limited to a specific subset of participants with schizophrenia rather than all diagnosed individuals. While there is still no consensus with regard to the exact stratification of SCZ with regard to the number and type of clusters based on cognitive or SC performance, previous studies have found at least one cognitively spared cluster of patients and either one or several clusters of patients with various levels of cognitive impairment (Green, Girshkin, Kremerskothen, Watkeys, & Quidé, 2020). As proposed by Ludwig et al. (2020), individuals with a psychotic disorder but “adequate social cognition and functioning may feel lonelier when experiencing a mismatch between their desired and current relationship quantity, quality or type” (p. 555). Thus, the lack of observed relationships between loneliness and SC in patients could stem from examining both constructs at the level of the whole clinical group instead of particular subgroups of patients differing in social cognitive capacity.

The aim of the current study is to explore social cognitive and neurocognitive mechanisms that may be associated with loneliness in schizophrenia through the process of mega-analysis (pooling raw data across studies) of two datasets, including data from the original Ludwig et al. (2020) study. First, we aim to re-examine the associations between demographic, clinical and cognitive variables, and loneliness in a large set of cross-national participants. Second, we test the "cluster hypothesis”, by investigating the association between social cognition and loneliness across subgroups of patients differing in SC capacity.

## Methods

### Participants

This study includes patients with a diagnosis of schizophrenia or schizoaffective disorder who participated in the original SCOPE study at the University of North Carolina site, which was the only SCOPE site that examined loneliness in participants (n=60; UNC; for details please see (Ludwig et al., 2020)). The second sample included in the mega-analysis comprised patients treated at the Institute of Psychiatry and Neurology in Warsaw who were examined as a part of the Polish National Centre of Science Sonata grant no 2016/23/D/HS6/02947 (SONATA; n=88). Adults (>18 years-old) diagnosed with schizophrenia according to ICD-10 criteria were recruited for the study. Only clinically stable patients who had not undergone any treatment changes and did not endorse any significant changes in symptom severity during the 4 weeks preceding the baseline study assessment visit were included in the study. Patients with intellectual disability, a history of comorbid neurological or psychiatric disorders, or drug abuse were excluded from the project.

Participants for the control group included healthy volunteers who responded to online advertisements. In both UNC and SONATA, healthy individuals were screened for exclusion criteria (history of psychiatric or neurological disorders, substance abuse), and groups were matched according to sex, age, and parental education. Each participant received written information about the study procedure and provided signed written consent before participating in the study. The study protocol was approved by the respective ethics boards at UNC and the Institute of Psychology, Polish Academy of Sciences. SONATA participants were reimbursed at the rate of 100 PLN (appx 25 USD), and UNC participants received 60 USD. The SONATA project had a more restrictive age criteria compared to the SCOPE study. As such, only participants aged 18-51 were included from the UNC sample into the current project.

For the pooled analysis, only individuals with no missing data for any of the variables of interest (loneliness score, PANSS scores, five neurocognitive tests and four SC measures) were included in the analyses. This produced a final sample of 251 individuals, including 148 SCZ (88 from SONATA) and 103 HC (65 from SONATA). One SCZ was excluded from further analysis due to extreme outlier values in TMT-A, thus leaving 250 participants for pooled analyses. The sample consisted of 160 males and 90 females; no gender differences were found between groups and sites (X^2^_1_=2.5; p=.14). No age differences were found between SCZ (34.30+/-8.61) and HC (32.40+/-8.16; F_1,249_=2.61; p=.11). Participants from UNC (34.76+/-8.39; n=97) were slightly older than participants from the SONATA sample (32.07+/-7.90; n=153; F_1,249)_= 11.29; p=.001). No interaction between group and site was found (F_1,249_<.01; p=.95). Patients also reported fewer years of education compared to controls (F_1,246_=25.23; p<.001). No between site differences were found for negative and cognitive PANSS scores, however patients from SONATA had fewer positive symptoms (F_1,146_=9.67; p=.002) and affective symptoms (F_1,146)_=25.19; p<.001) compared to patients from UNC. No difference in the CPZ equivalent or type of medication was found between the sites. Given the size of the SCZ group, the current study should be able to detect small-to-medium effects (Pearson’s r=.23 at p=.05 with 80% power), as estimated by *pwr* R library.

### Neurocognitive and SC tests

For the SONATA sample, the full Polish Academic version of MATRICS Consensus Cognitive Battery (MCCB) (Jędrasik-Styła et al., 2015) was used to examine cognitive functioning in participants. MCCB is a standardized battery consisting of ten tests, which assess seven cognitive domains (speed of processing, attention/vigilance, working memory, verbal learning, visual learning, reasoning, and problem-solving, social cognition). UNC participants completed only a subset of the MCCB, which included three tests measuring speed of processing (Trail Making Test – A, BACS Symbol Coding, Animal Fluency), one verbal learning task (Hopkins Verbal Learning Test – Revised) and one working memory test (Letter Number Span). For this reason, the current mega-analysis only included the five neurocognitive tests administered in both projects.

Four of the SC measures in the SCOPE battery are available in Polish, including two measures of mentalizing (Reading the Mind in the Eyes Task; RMET (Baron-Cohen, Wheelwright, Hill, Raste, & Plumb, 2001); Hinting Task (Krawczyk, Schudy, Jarkiewicz, & Okruszek, 2020)), a social perception task (the Mini Profile of Nonverbal Sensitivity; MiniPONS (Bänziger, Scherer, Hall, & Rosenthal, 2011)), and a test of emotion processing (Penn Emotion Recognition Task, PENN ER-40 (Gur et al., 2002)).

### Clinical assessment

Clinical symptom severity was assessed in both samples using the Positive and Negative Syndrome Scale (Kay & Opler, 1987). The four clinical domain scores (positive, negative, affective, cognitive) were calculated in line with recent PHAMOUS consortium recommendations, as it has been based on the large multinational sample and has been strongly linked to neurobiological markers (Chen et al., 2020). As the specific PANSS items N4 (Passive/apathetic social withdrawal) and G16 (Active social avoidance) are highly semantically linked to the constructs measured by loneliness questionnaires, the pattern of results was also re-investigated after dropping these two items from respective subscales.

### Loneliness measurement

Loneliness in the SONATA sample was measured using the Polish version of the Revised UCLA Loneliness scale (Kwiatkowska, Rogoza, & Kwiatkowska, 2017). The scale includes 20 statements which are reflective of one’s loneliness and has been shown to have excellent internal consistency when administered to a large group of non-clinical Polish participants (α=.92). Loneliness in UNC sample was examined using the initial version of the UCLA Loneliness Scale, which has been reworded to produce R-UCLA Loneliness scale. Considering the Likert scale for responses on the initial version of the UCLA scale range from 0 to 3, instead of 1 to 4 as is the case for the updated R-UCLA scale, total scores for the UNC sample have been shifted by adding 20 points to the overall score for participants from UNC to produce a common scale for both samples.

### Statistical analysis

Data analyses were performed using Statistical Package for the Social Sciences (SPSS 27) and R libraries. We completed a two-factor ANOVA with site (Poland vs USA) and group (SCZ vs HC) as between-subject factors to examine differences in neurocognitive and SC variables. Regarding loneliness, an additional factor (gender) was also investigated, as it has been significantly associated with loneliness in the original Ludwig et al. (2020) analysis. To examine the relationship between age, clinical, and cognitive variables and loneliness, Pearson correlation coefficients have been calculated separately for SCZ and HC.

Next, Latent Class Analysis (LCA) was implemented using Mclust (version 5.4.7) library to identify latent classes of patients that present similar SC profiles (Scrucca, Fop, Murphy, & Raftery, 2016). Mclust provides functions for parameter estimation using the expectation-maximization algorithm for normal mixture models. One of the main advantages of using Mclust is the use of the Bayesian information criterion (BIC) which selects the optimal covariance structure and number of clusters. Furthermore, LCA has been demonstrated to produce fewer misclassifications compared to k-means clustering (Magidson & Vermunt, 2002). LCA classification was produced on the basis of the patients’ performance on four SC tests (Mini-PONS, ER-40, RMET, Hinting) that were entered into the clustering algorithm. As most of the studies observed non-impaired cognitive performance in at least one cluster of patients, SC scores from LCA clusters were compared with HC. Finally, the pattern of correlations between cognitive variables and loneliness were reinspected separately for each LCA cluster.

## Results

### Neurocognitive and SC tests

In the first step of the analysis, we have examined the factors associated with neurocognitive and social cognitive scores in the current study. Patients performed worse than HC on all assessments (effect sizes ranging from d=0.30 to d=1.23, Table 1). Main effects of site were found for the UNC sample demonstrating worse performance on Trails A (d=0.28), animal fluency (d=0.49), HVLT-R (d=0.32), and Hinting Task (d=0.73), but better performance for Mini-PONS (d=0.28). However, differences became non-significant after age was entered as a covariate on TMT and HVLT-R. No group by site interactions were observed for any of the tests.

**Table 1.** Between-group differences for neurocognitive and social cognitive tasks.

|  | SCZ (n=147) | HC (n=103) | Cohen's d and statistics |
| --- | --- | --- | --- |
| TMT-A (time) | 35.92 (13.23) | 28.14 (8.59) | d=.67 p<.001 $F_{1,249}=26.73$ |
| BACS (coded symbols) | 45.55 (10.82) | 59.29 (11.73) | d=1.23 p<.001 $F_{1,249}=85.35$ |
| HVLT-R (words) | 24.08 (5.48) | 26.95 (4.52) | d=.56 p<.001 $F_{1,249}=17.35$ |
| LNS (points) | 13.67 (3.72) | 15.36 (3.02) | d=.49 p<.001 $F_{1,249}=13.12$ |
| Animal Fluency (no of items) | 22.52 (6.03) | 27.41 (5.93) | d=.81 p<.001 $F_{1,249}=35.60$ |
| PENN ER-40 (correct answers) | 32.00 (3.92) | 33.04 (2.80) | d=.30 p<.05 $F_{1,249}=5.87$ |
| PONS (correct answers) | 44.13 (5.37) | 47.80 (5.40) | d=.68 p<.001 $F_{1,249}=27.93$ |
| HT (points) | 14.46 (3.07) | 16.44 (2.64) | d=.68 p<.001 $F_{1,249}=25.26$ |
| RMET (correct answers) | 23.68 (5.06) | 26.02 (3.87) | d=.51 p<.001 $F_{1,249}=12.95$ |

### Loneliness

In the second step of analysis, we examined the factors associated with loneliness in the current study. SCZ reported higher levels of loneliness (44.31+/-12.73) compared to HC (34.17+/-10.23; F_1,249_=52.20; p<.001; d=0.86). This effect was not moderated by gender. A weak group by site interaction was found (F_1,249_=5.82; p=.017) such that in each sample, SCZ were lonelier than HC, however UNC HC were less lonely compared to SONATA HC, while the reverse pattern was observed in SCZ. Furthermore, no correlation between age and loneliness was found in SCZ (r=.06; p=.47), whereas these variables were negatively correlated in HC (r=-.22; p=.02). A robust correlation was found for loneliness with PANSS negative (r=.25; p=.002) and affective (r=.49 p<.001) dimensions and a trend-level correlation with positive symptoms (r=.15; p=.08). The same pattern of results remained even after excluding N4 (Passive/apathetic social withdrawal) and G16 (Active social avoidance) from PHAMOUS Negative and Affective dimensions, respectively.

### Association between loneliness and cognitive variables in groups

During the final stage of the whole-group analyses, we examined the association between neurocognitive and social cognitive scores and loneliness at the level of whole groups. There were no significant relationships between loneliness and any of the cognitive variables within the full SCZ group (r from -.10 to .14). In HC, loneliness was negatively associated with performance on the Mini-PONS (r=-.22; p<.05; Table 2).

**Table 2.** Zero-order correlations between loneliness and neurocognitive and social cognitive tasks.

|  | SCZ (n=147) | HC (n=103) |
| --- | --- | --- |
| RMET | .137 | -.043 |
| PENN ER-40 | -.095 | -.099 |
| HT | -.010 | .152 |
| PONSS | .076 | -.221* |
| HVLT-R | -.008 | -.075 |
| TMT-A | .142 | .095 |
| BACS | -.046 | -.154 |
| Animal Fluency | -.016 | -.077 |
| LNS | .064 | -.048 |

### SC clusters

Before examining the “cluster hypothesis”, the results of the LCA were investigated. The lowest BIC was observed for the diagonal model assuming unequal volume, but equal shape of the clusters (model “VEI”; Figure 1). Each of the top three models based on the BIC criterion had two clusters, thus the VEI two-cluster solution with 65 and 82 patients, respectively, was further investigated. Clusters did not differ in age (t_145_=.18; p=.86; d=0.03), gender (females: 16/65 vs. 31/82; X^2^_1_=.3; p=.11), percentage of participants from each site (UNC participants: 29/65 vs. 30/82; X^2^_1_=1.0; p=.40), or loneliness (t_145_=1.09; p=.28; d=0.18). Finally, no between-clusters differences were observed for PHAMOUS positive (t_145_=1.55; p=.12; d=0.26), negative (t_145_=.58; p=.57; d=0.10) or affective (t_145_=.04; p=.97; d<0.01) symptoms. The smaller cluster, however, had more cognitive symptoms compared to the larger one (t_145_=2.42; p=.02; d=0.40). Within the UNC sample a higher percentage of non-Caucasian participants was found in the smaller cluster (18 out of 29) compared to the larger cluster (4 out of 30; X^2^_4_=15.4; p=.004). For each of the four SC tests, the same pattern of results was found, confirming previous findings suggesting the existence of two clusters differing in social cognitive capacity in patients with schizophrenia: patients fell into either a normal-performance group (SC-NP; n=82) that did not differ from HC, or a SC-impaired group (SC-IMP, n=65) who performed worse on all four measures compared both to SC-NP and HC (all *p*s<.001).

**Figure 1.**
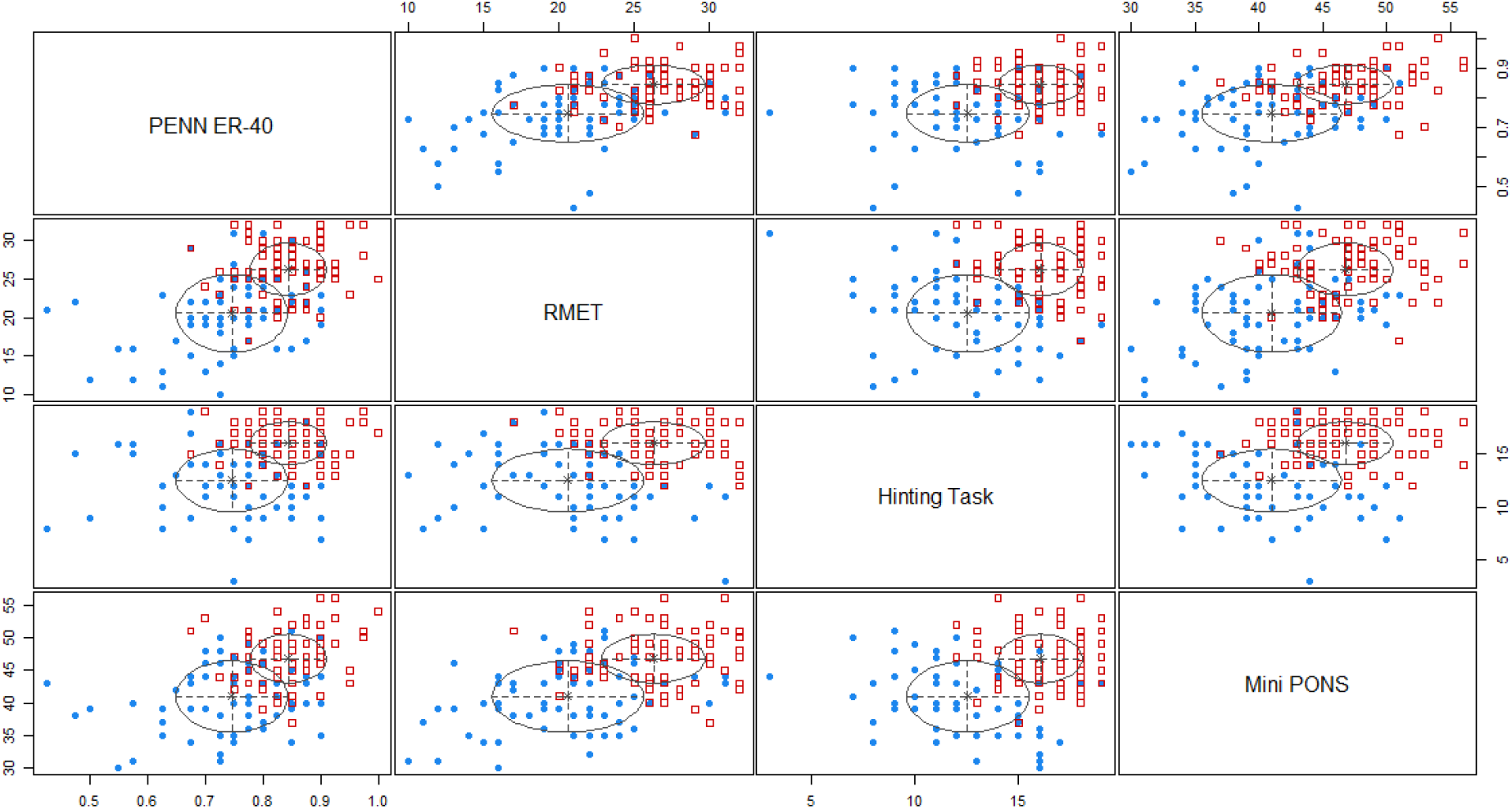
Two-cluster classification produced with Latent Class Analysis.

### Association between loneliness and cognitive variables in SC clusters

In a last step of analysis, the “cluster hypothesis” was evaluated by examining the association between neurocognitive and social cognitive variables and loneliness separately in SC-NP and SC-IMP clusters. No correlations between loneliness and social cognitive or neurocognitive scores were observed for the SC-NP cluster. However, in the SC-IMP cluster a negative association was found between loneliness and mentalizing abilities (Hinting: r=-.25; p<.05). Moreover, trend-level associations were found for emotion-recognition (PENN ER-40: r=-.22; p<.08) and speed of processing (TMT-A: r=.22; p<.08) with loneliness. To further evaluate the independent and combined contributions of neurocognitive and social cognitive processes on loneliness in the SC-IMP cluster, the three variables were entered in subsequent steps of a regression model. Mentalizing abilities explained 5% of the loneliness in the SC-IMP cluster (adjusted R^2^=4.7%; F_1,63_=4.12; p<.05). Emotion recognition skills entered after mentalizing skills contributed an additional 4% of variance in loneliness scores (R^2^-change=4.4%; p<.05). The incremental increase in variance for TMT was not significant (R^2^-change=1.2%; p=.18).

## Discussion

The current study extends previous knowledge about the factors which may underlie loneliness in patients with schizophrenia. First, in line with previous studies, we observed increased levels of loneliness reported by individuals with schizophrenia compared to individuals without the disorder. In fact, the magnitude of the effect was similar to that which was reported in the meta-analysis of thirteen studies on the association between psychotic symptoms and loneliness (Michalska da Rocha et al., 2018). This effect was not modulated by age or gender of patients and was consistently observed in both samples from different cultures included in the current mega-analysis, thus suggesting that loneliness may be seen as a universal problem across SCZ. Additionally, it is worth noting that none of the between-group differences observed in SC tasks was moderated by the site of the study. Lack of cross-cultural validation has been recently listed among the main challenges in research on SC impairment in SCZ (Vaskinn & Horan, 2020). Thus, the similarity of findings across Polish and American participants in the current study adds to the previous results suggesting stability of findings with SCOPE battery across cultures (Lim, Lee, Pinkham, Lam, & Lee, 2020).

Second, similar to previous studies demonstrating a robust association between depression and loneliness in both the general population (Cacioppo, Hughes, Waite, Hawkley, & Thisted, 2006) and in individuals with mental health crises (Wang et al., 2020), we have found that more severe affective symptoms were the most robust clinical predictor of loneliness in patients. However, it is worth noting that chronic loneliness has been shown to increase the risk of depressive symptoms in healthy individuals, but the reverse relationship was not found (Cacioppo, Hawkley, & Thisted, 2010). As the cross-sectional design of the current study precludes establishing the direction of the causality between affective symptoms and loneliness in patients, further longitudinal investigation is needed to establish whether this observation applies to SCZ. Given that negative symptoms are often considered as closely related with social dysfunction in schizophrenia (Robertson et al., 2014), it is notable that we observed a stable and small-to-medium relationship between negative symptoms and loneliness in patients.

Importantly, we did not observe an association between SC capacity and loneliness in the whole sample of patients. The size of the current sample was large enough to detect small-to-medium correlations, which confirms that previous null-findings in smaller samples (e.g. 74 patients reported by Ludwig et al. (2020) or 87 patients included in Trémeau et al. (2016)) were not due to insufficient statistical power. In contrast to the two previous studies, we employed a data-driven latent profile analysis of SC performance to stratify the SCZ sample into clusters differing in SC-capacity. We identified a two-cluster structure, with the normal-performance cluster of patients outperforming the SC-impaired cluster on all neuro- and social cognitive tasks. Importantly, no between-group differences across key demographic characteristics or loneliness levels were observed. Yet, while no association was found between loneliness and SC in unimpaired groups, poorer theory of mind and emotion recognition was linked to higher levels of loneliness in the SC-impaired sample.

This finding is congruent with Ludwig et al.’s notion that the association between social cognitive ability and loneliness may be limited to a specific subgroup of SCZ. However, while Ludwig et al. (2020) proposed that patients with "adequate” social cognition and functioning may feel lonelier as they are better suited to appraise the mismatch between their desired and actual social functioning, we have observed no significant relationship between SC and loneliness in SC-NP sample. This finding is similar to the previous findings from healthy individuals (Okruszek et al., 2021). Moreover, a recent review concluded that problems with understanding and interpreting social situations in lonely individuals may be attributed to negative cognitive bias rather than perceptual or attentional deficits (Spithoven et al., 2017). Thus, it may be suggested that in individuals with intact social cognition (whether HC or SC-NP), loneliness may be associated with SC tendencies (e.g. hostile attribution bias (Okruszek et al., 2021)) rather than SC capacity per se. At the same time, previous findings suggest a negative association between SC capacity and loneliness in other cognitively-impaired samples, e.g. stroke patients (Adams et al., 2020). Accordingly, the current study found that the relationship between loneliness and SC capacity may be limited to the individuals with schizophrenia with SC impairments.

Taken together, it may be proposed that in individuals with unimpaired SC, more subtle mechanisms associated with tendencies to interpret social situations in a negative way may be associated with loneliness, while relationship between loneliness and baseline SC capacity is nonsignificant. However, after reaching a SC-deficit threshold, overall SC capacity becomes crucial for one’s level of loneliness, e.g., with less impaired SC serving as a buffer against loneliness in SC-impaired participants. Consistent with precision psychiatry postulates (Williams, 2016), the findings of the current study stress the importance of embracing social cognitive heterogeneity in patients with schizophrenia while developing psychosocial interventions targeting loneliness in SCZ. A recent large scale study focused solely on social cognitive processes found a significant difference in the outcome and clinical variables between clusters demonstrating varied SC impairment (Rocca et al., 2016). Similarly, decreased social functioning was found in a poorly-performing cluster in a study which utilized neuroimaging data for biophenotyping (Viviano et al., 2018). Here, we extend these findings by showing that even in the absence of differences in loneliness between normal-performing and SC-impaired clusters, different mechanisms may be associated with loneliness in each cluster of SCZ. Thus, planning individualized treatment needs to consider both general levels of SC capacity and specific profiles of SC abilities in each patient.

While the current set of analyses was well-powered and included a wide range of measures, some limitations of the current design should be noted. First, the cross-sectional design limits the ability to infer which factors may be predictors or consequences of loneliness in persons with schizophrenia. Second, because the current study merged data from two different sites, it is important to note that two different versions of the UCLA loneliness scale were administered in each study. Despite the consistency of findings across our samples which suggests robustness and replicability, future research with congruent methodology should be considered. Furthermore while we utilized an extensive battery to investigate SC capacity, future studies should also include assessment of factors linked to social motivation and cognitive biases which may also be of great importance for loneliness in SCZ (Green et al., 2018), especially given the fact that the SC predictors accounted only for 9% of loneliness variance in the SCZ-IMP cluster.

## Data Availability

All data produced in the present study are available upon reasonable request to the authors

## Acknowledgments

This research was supported by Grant MH93432 to Drs. Harvey, Penn, and Pinkham from the National Institute of Mental Health, and by Grant 2016/23/D/HS6/02947 to Dr Okruszek from the Polish National Centre of Science.

## Bibliography

Adams, A. G., Henry, J. D., Molenberghs, P., Robinson, G. A., Nott, Z., & von Hippel, W. (2020). The relationship between social cognitive difficulties in the acute stages of stroke and later functional outcomes. Social Neuroscience, 15(2), 158–169. doi:10.1080/17470919.2019.1668845

Badcock, J. C., Mackinnon, A., Waterreus, A., Watts, G. F., Castle, D., McGrath, J. J., & Morgan, V. A. (2019). Loneliness in psychotic illness and its association with cardiometabolic disorders. Schizophrenia Research, 204, 90–95. doi:10.1016/j.schres.2018.09.021

Badcock, J. C., Shah, S., Mackinnon, A., Stain, H. J., Galletly, C., Jablensky, A., & Morgan, V. A. (2015). Loneliness in psychotic disorders and its association with cognitive function and symptom profile. Schizophrenia Research, 169(1-3), 268–273. doi:10.1016/j.schres.2015.10.027

Bänziger, T., Scherer, K. R., Hall, J. A., & Rosenthal, R. (2011). Introducing the minipons: A short multichannel version of the profile of nonverbal sensitivity (PONS). Journal of nonverbal behavior, 35(3), 189–204. doi:10.1007/s10919-011-0108-3

Baron-Cohen, S., Wheelwright, S., Hill, J., Raste, Y., & Plumb, I. (2001). The “Reading the Mind in the Eyes” Test revised version: a study with normal adults, and adults with Asperger syndrome or high-functioning autism. Journal of Child Psychology and Psychiatry, and Allied Disciplines, 42(2), 241–251. doi:10.1017/S0021963001006643

Cacioppo, J. T., & Cacioppo, S. (2018). Loneliness in the modern age: an evolutionary theory of loneliness (ETL) (Vol. 58, pp. 127–197). Elsevier. doi:10.1016/bs.aesp.2018.03.003

Cacioppo, J. T., Hawkley, L. C., & Thisted, R. A. (2010). Perceived social isolation makes me sad: 5-year cross-lagged analyses of loneliness and depressive symptomatology in the Chicago Health, Aging, and Social Relations Study. Psychology and Aging, 25(2), 453–463. doi:10.1037/a0017216

Cacioppo, J. T., Hughes, M. E., Waite, L. J., Hawkley, L. C., & Thisted, R. A. (2006). Loneliness as a specific risk factor for depressive symptoms: cross-sectional and longitudinal analyses. Psychology and Aging, 21(1), 140–151. doi:10.1037/0882-7974.21.1.140

Chau, A. K. C., Zhu, C., & So, S. H.-W. (2019). Loneliness and the psychosis continuum: a meta-analysis on positive psychotic experiences and a meta-analysis on negative psychotic experiences. International Review of Psychiatry, 31(5-6), 471–490. doi:10.1080/09540261.2019.1636005

Chen, J., Patil, K. R., Weis, S., Sim, K., Nickl-Jockschat, T., Zhou, J., … Pharmacotherapy Monitoring and Outcome Survey (PHAMOUS) Investigators. (2020). Neurobiological Divergence of the Positive and Negative Schizophrenia Subtypes Identified on a New Factor Structure of Psychopathology Using Non-negative Factorization: An International Machine Learning Study. Biological Psychiatry, 87(3), 282–293. doi:10.1016/j.biopsych.2019.08.031

Eglit, G. M. L., Palmer, B. W., Martin, A. S., Tu, X., & Jeste, D. V. (2018). Loneliness in schizophrenia: Construct clarification, measurement, and clinical relevance. Plos One, 13(3), e0194021. doi:10.1371/journal.pone.0194021

Green, M. F., Horan, W. P., Lee, J., McCleery, A., Reddy, L. F., & Wynn, J. K. (2018). Social disconnection in schizophrenia and the general community. Schizophrenia Bulletin, 44(2), 242–249. doi:10.1093/schbul/sbx082

Green, M. J., Girshkin, L., Kremerskothen, K., Watkeys, O., & Quidé, Y. (2020). A Systematic Review of Studies Reporting Data-Driven Cognitive Subtypes across the Psychosis Spectrum. Neuropsychology Review, 30(4), 446–460. doi:10.1007/s11065-019-09422-7

Gur, R. C., Sara, R., Hagendoorn, M., Marom, O., Hughett, P., Macy, L., … Gur, R. E. (2002). A method for obtaining 3-dimensional facial expressions and its standardization for use in neurocognitive studies. Journal of Neuroscience Methods, 115(2), 137–143. doi:10.1016/s0165-0270(02)00006-7

Halverson, T., Orleans-Pobee, M., Merritt, C., Sheeran, P., Fett, A. K., & Penn, D. L. (2019). Pathways to functional outcomes in schizophrenia spectrum disorders: Meta-analysis of social cognitive and neurocognitive predictors. Neuroscience and Biobehavioral Reviews, 105, 212–219. doi:10.1016/j.neubiorev.2019.07.020

Holt-Lunstad, J., Smith, T. B., Baker, M., Harris, T., & Stephenson, D. (2015). Loneliness and social isolation as risk factors for mortality: a meta-analytic review. Perspectives on Psychological Science, 10(2), 227–237. doi:10.1177/1745691614568352

Jędrasik-Styła, M., Ciołkiewicz, A., Styła, R., Linke, M., Parnowska, D., Gruszka, A., … Wichniak, A. (2015). The polish academic version of the MATRICS consensus cognitive battery (MCCB): evaluation of psychometric properties. The Psychiatric Quarterly, 86(3), 435–447. doi:10.1007/s11126-015-9343-9

Kay, S. R., & Opler, L. A. (1987). The positive-negative dimension in schizophrenia: its validity and significance. Psychiatric developments, 5(2), 79–103.

Krawczyk, M. M., Schudy, A. M., Jarkiewicz, M., & Okruszek, Ł. (2020). Polish version of the Hinting Task - pilot study with patients with schizophrenia. Psychiatria Polska, 54(4), 727–739. doi:10.12740/PP/112265

Kwiatkowska, M. M., Rogoza, R., & Kwiatkowska, K. (2017). Analysis of the psychometric properties of the Revised UCLA Loneliness Scale in a Polish adolescent sample. Current Issues in Personality Psychology, 6(2), 164–170. doi:10.5114/cipp.2017.69681

Lim, K., Lee, S.-A., Pinkham, A. E., Lam, M., & Lee, J. (2020). Evaluation of social cognitive measures in an Asian schizophrenia sample. Schizophrenia research. Cognition, 20, 100169. doi:10.1016/j.scog.2019.100169

Lim, M. H., Gleeson, J. F. M., Alvarez-Jimenez, M., & Penn, D. L. (2018). Loneliness in psychosis: a systematic review. Social Psychiatry and Psychiatric Epidemiology, 53(3), 221–238. doi:10.1007/s00127-018-1482-5

Ludwig, K. A., Nye, L. N., Simmons, G. L., Jarskog, L. F., Pinkham, A. E., Harvey, P. D., & Penn, D. L. (2020). Correlates of loneliness among persons with psychotic disorders. Social Psychiatry and Psychiatric Epidemiology, 55(5), 549–559. doi:10.1007/s00127-019-01789-5

Magidson, J., & Vermunt, J. (2002). Latent class models for clustering: A comparison with K-means. Canadian journal of marketing research.

Michalska da Rocha, B., Rhodes, S., Vasilopoulou, E., & Hutton, P. (2018). Loneliness in Psychosis: A Meta-analytical Review. Schizophrenia Bulletin, 44(1), 114–125. doi:10.1093/schbul/sbx036

Okruszek, Ł., Piejka, A., Krawczyk, M., Schudy, A., Wiśniewska, M., Żurek, K., & Pinkham, A. (2021). Owner of a lonely mind? Social cognitive capacity is associated with objective, but not perceived social isolation in healthy individuals. Journal of research in personality, 104103. doi:10.1016/j.jrp.2021.104103

Pinkham, A. E., Brensinger, C., Kohler, C., Gur, R. E., & Gur, R. C. (2011). Actively paranoid patients with schizophrenia over attribute anger to neutral faces. Schizophrenia Research, 125(2-3), 174–178. doi:10.1016/j.schres.2010.11.006

Ringen, P. A., Engh, J. A., Birkenaes, A. B., Dieset, I., & Andreassen, O. A. (2014). Increased mortality in schizophrenia due to cardiovascular disease - a non-systematic review of epidemiology, possible causes, and interventions. Frontiers in psychiatry, 5, 137. doi:10.3389/fpsyt.2014.00137

Robertson, B. R., Prestia, D., Twamley, E. W., Patterson, T. L., Bowie, C. R., & Harvey, P. D. (2014). Social competence versus negative symptoms as predictors of real world social functioning in schizophrenia. Schizophrenia Research, 160(1-3), 136–141. doi:10.1016/j.schres.2014.10.037

Rocca, P., Montemagni, C., Mingrone, C., Crivelli, B., Sigaudo, M., & Bogetto, F. (2016). A cluster-analytical approach toward real-world outcome in outpatients with stable schizophrenia. European Psychiatry, 32, 48–54. doi:10.1016/j.eurpsy.2015.11.007

Russell, D., Peplau, L. A., & Cutrona, C. E. (1980). The revised UCLA Loneliness Scale: concurrent and discriminant validity evidence. Journal of Personality and Social Psychology, 39(3), 472–480. doi:10.1037/0022-3514.39.3.472

Schneider, M., Reininghaus, U., van Nierop, M., Janssens, M., Myin-Germeys, I., & GROUP Investigators. (2017). Does the Social Functioning Scale reflect real-life social functioning? An experience sampling study in patients with a non-affective psychotic disorder and healthy control individuals. Psychological Medicine, 47(16), 2777–2786. doi:10.1017/S0033291717001295

Scrucca, L., Fop, M., Murphy, T. B., & Raftery, A. E. (2016). mclust 5: Clustering, Classification and Density Estimation Using Gaussian Finite Mixture Models. The R journal, 8(1), 289–317. doi:10.32614/RJ-2016-021

Siegrist, K., Millier, A., Amri, I., Aballéa, S., & Toumi, M. (2015). Association between social contact frequency and negative symptoms, psychosocial functioning and quality of life in patients with schizophrenia. Psychiatry Research, 230(3), 860–866. doi:10.1016/j.psychres.2015.11.039

Spithoven, A. W. M., Bijttebier, P., & Goossens, L. (2017). It is all in their mind: A review on information processing bias in lonely individuals. Clinical Psychology Review, 58, 97–114. doi:10.1016/j.cpr.2017.10.003

Stain, H. J., Galletly, C. A., Clark, S., Wilson, J., Killen, E. A., Anthes, L., … Harvey, C. (2012). Understanding the social costs of psychosis: the experience of adults affected by psychosis identified within the second Australian National Survey of Psychosis. The Australian and New Zealand Journal of Psychiatry, 46(9), 879–889. doi:10.1177/0004867412449060

Trémeau, F., Antonius, D., Malaspina, D., Goff, D. C., & Javitt, D. C. (2016). Loneliness in schizophrenia and its possible correlates. An exploratory study. Psychiatry Research, 246, 211–217. doi:10.1016/j.psychres.2016.09.043

Vaskinn, A., & Horan, W. P. (2020). Social cognition and schizophrenia: unresolved issues and new challenges in a maturing field of research. Schizophrenia Bulletin, 46(3), 464–470. doi:10.1093/schbul/sbaa034

Viviano, J. D., Buchanan, R. W., Calarco, N., Gold, J. M., Foussias, G., Bhagwat, N., … Social Processes Initiative in Neurobiology of the Schizophrenia(s) Group. (2018). Resting-State Connectivity Biomarkers of Cognitive Performance and Social Function in Individuals With Schizophrenia Spectrum Disorder and Healthy Control Subjects. Biological Psychiatry, 84(9), 665–674. doi:10.1016/j.biopsych.2018.03.013

Wang, J., Lloyd-Evans, B., Marston, L., Ma, R., Mann, F., Solmi, F., & Johnson, S. (2020). Epidemiology of loneliness in a cohort of UK mental health community crisis service users. Social Psychiatry and Psychiatric Epidemiology, 55(7), 811–822. doi:10.1007/s00127-019-01734-6

Williams, L. M. (2016). Precision psychiatry: a neural circuit taxonomy for depression and anxiety. The Lancet. Psychiatry, 3(5), 472–480. doi:10.1016/S2215-0366(15)00579-9

